# Deep phenotyping of patient lived experience in functional bowel disorders

**DOI:** 10.1101/2024.01.23.24301624

**Authors:** James K. Ruffle, Michelle Henderson, Cho Ee Ng, Trevor Liddle, Amy P. K. Nelson, Parashkev Nachev, Charles H Knowles, Yan Yiannakou

**Author notes:** Correspondence to: Dr James K Ruffle Address: Institute of Neurology, UCL, London, UK. Correspondence may also be addressed to: Professor Yan Yiannakou Address: Durham Bowel Dysfunction Service, University Hospital North Durham, County Durham and Darlington NHS Trust, Durham, UK.

## Abstract

Clinical management relies on a diagnostic label as the primary guide to treatment. However, individual patients’ lived experiences vary more widely than standard diagnostic categories reflect. This is especially true for functional bowel disorders (FBDs), a heterogeneous and challenging group of gastrointestinal disorders where no definitive diagnostic tests, clinical biomarkers, or universally effective treatments exist. Characterising the link between disease and lived experience - in the face of marked patient heterogeneity - requires deep phenotyping of the interactions between multiple characteristics plausibly achievable only with complex modelling approaches. In a large patient cohort (n=1175), we developed a machine learning and Bayesian generative graph framework to better understand the lived experience of FBDs. Iterating through 59 factors available from routine clinical care, spanning patient demography, diagnosis, symptomatology, life impact, mental health indices, healthcare access requirements, COVID-19 impact, and treatment effectiveness, machine models were used to quantify the predictive fidelity of one feature from the remainder. Bayesian stochastic block models were used to delineate the network community structure underpinning the heterogeneous lived experience of FBDs. Machine models quantified patient personal health rating (R^2^ 0.35), anxiety and depression severity (R^2^ 0.54), employment status (balanced accuracy 96%), frequency of healthcare attendance (R^2^ 0.71), and patient-reported treatment effectiveness variably (R^2^ range 0.08-0.41). Contrary to the view of many healthcare professionals, the greatest determinants of patient-reported health and quality-of-life were life impact, mental well-being, employment status, and age, rather than diagnostic group and symptom severity. Patients responsive to one treatment were more likely to respond to another, leaving many others refractory to all. Clinical assessment of patients with FBDs should be less concerned with diagnostic classification than with the wider life impact of illness, including mental health and employment. The stratification of treatment response (and resistance) has implications for clinical practice and trial design, in need of further research.

## Introduction

Nosology is the branch of medical science that seeks to assign a disease label to a given pathological state. This is fundamentally a classification task where clinical history, examination, and investigational (such as serology or imaging) features are used to reach a diagnosis. However, there is much more to the individual patient’s experience than the disease label, to which a purely nosological classification is limited. Yet in contemporary medical research—and clinical practice—an objective characterisation of this aspect is rare.

A crucial factor in every disease and every individual patient is the patient’s lived experience^1^: the knowledge and understanding from personally living through something^2^. Clinically, lived experience is typically conceived as merely downstream of critical diagnostic features. But in many disorders, the patient’s lived experience is tightly interwoven with the diagnosis. Understanding a patient’s lived experience therefore merits the closest investigation across all available experiential, physiological, and pathophysiological levels.

Functional bowel disorders (FBDs) are a key example here^3,4^. Lived experience (often incorporating many years ahead of a diagnosis being reached^5^) is deeply embedded in diagnostic symptomatology. These are an exceptionally complex group of diseases with multiple biopsychosocial aspects, rendering them unique to each affected individual. They differ radically from most other gastrointestinal (GI) disorders, where clinical assessment and/or an investigational test can be used to assign a disease label and prescribe a treatment algorithm, and this is arguably a key reason that diagnostic and therapeutic innovation for FBDs has been considerably slower in progress.

Managing patients with FBDs is challenging for many reasons, including marked heterogeneity both across patients and in time for an individual, the absence of precise diagnostic tests and clinical biomarkers, and a diagnostic classification based on symptom profiles that may overlap and change^6^. An incomplete understanding of FBD pathophysiology has held back the development of targeted treatments^7^, including the prediction of which patients stand to benefit most from them, leaving this affected group of patients inequitably disadvantaged compared with many other disease groups.

The healthcare professional’s diagnostic classification of FBDs is based on varying combinations of GI symptoms. A patient’s experience of FBDs, however, is governed not merely by perturbation of the gastrointestinal tract but has far-reaching impact across the broader aspect of their life, ranging from the effect upon daily activities, mental well-being, access and satisfaction of healthcare, and treatment efficacy^3,4,6,8-11^. Patients affected by FBDs typically remain so for many years; living with these disorders becomes a necessity, leading to a considerable impact on quality of life (QoL), often with deleterious effects on other aspects of health^8,12^.

While FBDs are multifaceted high-dimensional disorders interwoven with the lived experience, they – like most other diseases - are rarely statistically modelled as such^6^. Clinical research studies often investigate these syndromes within relatively low-dimensional and/or linear statistical frameworks: common experimental designs may, for example, explore sex differences between disorder *x*, or age-related effects of treatment *y*, but rarely provide a more comprehensive investigation of multiple factors and their interactions. Such approaches invariably neglect many factors that define the individual, leaving gaps in our understanding of both the patient’s disorder and their lived experience^1^. The same applies to contemporary quantitative lived experience measures, such as quality of life (QOL) questionnaires, where a complex and multivariate pattern is compressed into a univariate score. Whilst such an approach is highly accessible, explaining its broad appeal across healthcare, more complex models offer substantially greater insight into the whole patient experience, with correspondingly greater actionable and translational potential.

The task requires deep phenotyping of the patient’s lived experience that has yet to be attempted at the scale and expressivity FBDs demand. We therefore developed a comprehensive pipeline harnessing state-of-the-art machine learning and Bayesian generative graph models to characterise, in unprecedented detail, the patient lived experience, prototyping it across a large UK cohort of patients with FBDs. By placing the individual patient’s perspective at the forefront of our approach^1^, we reveal the determinants of ill health, such as the impact on QoL and treatment effectiveness, in a more meaningful and patient-orientated way. This framework could pave the way to more richly individualised patient care^1,13-15^.

## Results

### Cohort

We received 1175 responses from 4739 patients (response rate 24.8%), who formed our analysis cohort. The mean age was 52 years (range 20-80 years): female (n=1000) and male (n=175) (Figure 1). This sex distribution aligns with existing research^16^. 642 patients fulfilled the ROME-IV criteria^3^ for IBS (IBS-C n=133, IBS-M n=237 IBS-D n=246, IBS-U n=26). Further functional bowel disorder diagnoses yielded were functional constipation (n=173) and functional diarrhoea (n=157). The remaining 203 patients demonstrated symptoms rendering them non-classifiable due to syndromic overlap in (n=130), or exclusion from (n=73), current classification systems.

**Figure 1.**
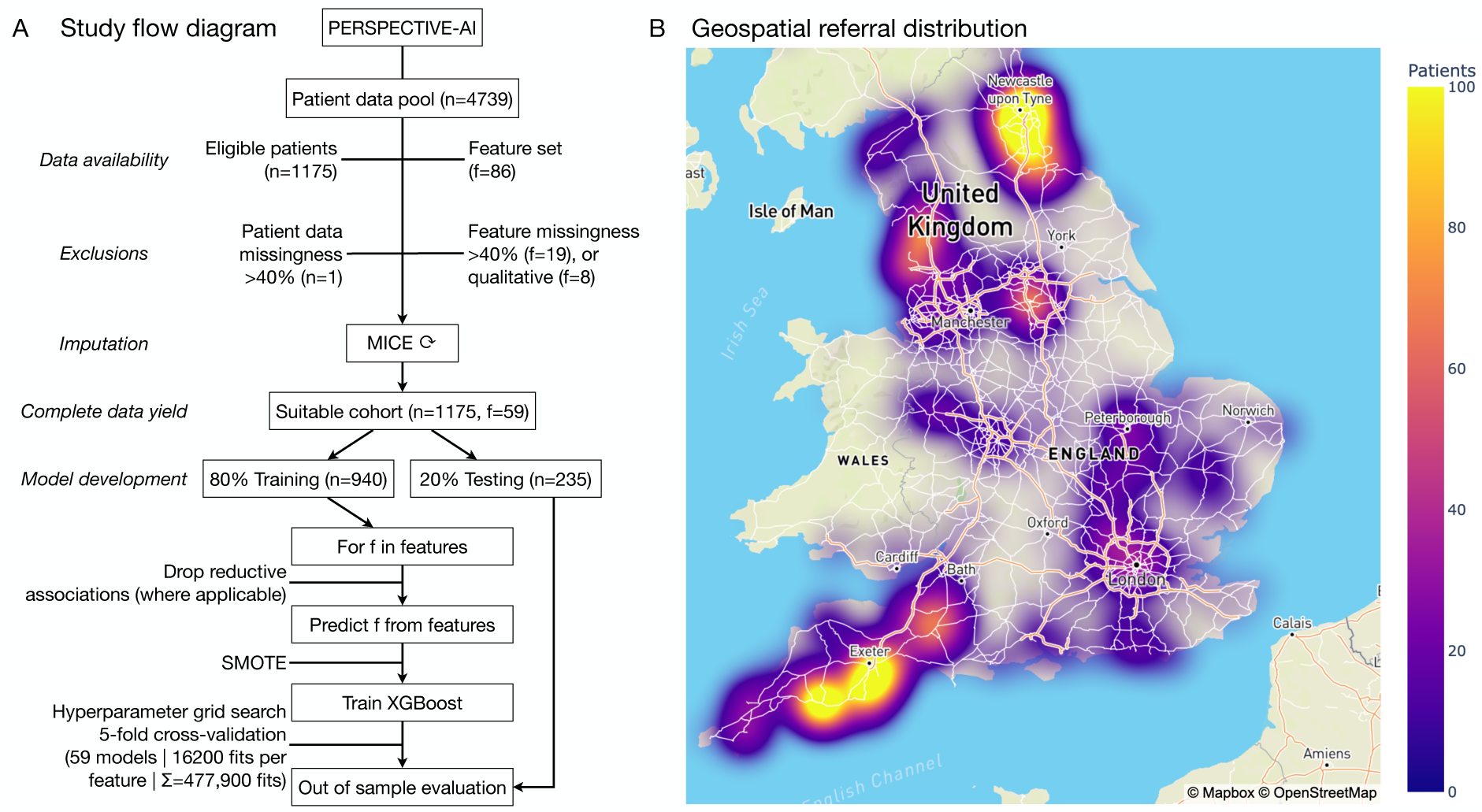
Study design. A) Flow diagram. B) Geospatial referral distribution.

We derived a hierarchical clustering representation of how patient factors were interrelated by pairwise correlation coefficient (Figure 2). This illustrated that when comparing simplistic linear relationships between patient factors, they generally align to self-explanatory domains. For example, measures of irrigation treatment use and patient-reported effectiveness were highly correlated and clustered together (r 0.64 or higher, all FDR-corrected p<0.0001). Similarly, measures of pain were highly correlated and clustered together, as well as pain-criterion diagnoses such as IBS (r 0.40 or higher, all FDR-corrected p<0.0001). Patient-reported effectiveness of several treatments formed another cluster, both medicinal (laxative use) and non-medicinal (including changes to diet, fluid intake, footstall use, and pelvic or sphincter exercises) (r range 0.13-0.50, all FDR-corrected p<0.0001). Pain severity, the impact of bowel symptoms on daily activities, and the impact on measures of assisted daily living (ADLs) formed another cluster (r 0.32 or higher, all FDR-corrected p<0.0001). Finally, engagement and requirement of healthcare services formed a weak cluster, including if seen by a medical consultant, general practitioner, dietician, and nurse (r range 0.12-0.45, FDR-corrected p<0.0001).

**Figure 2.**
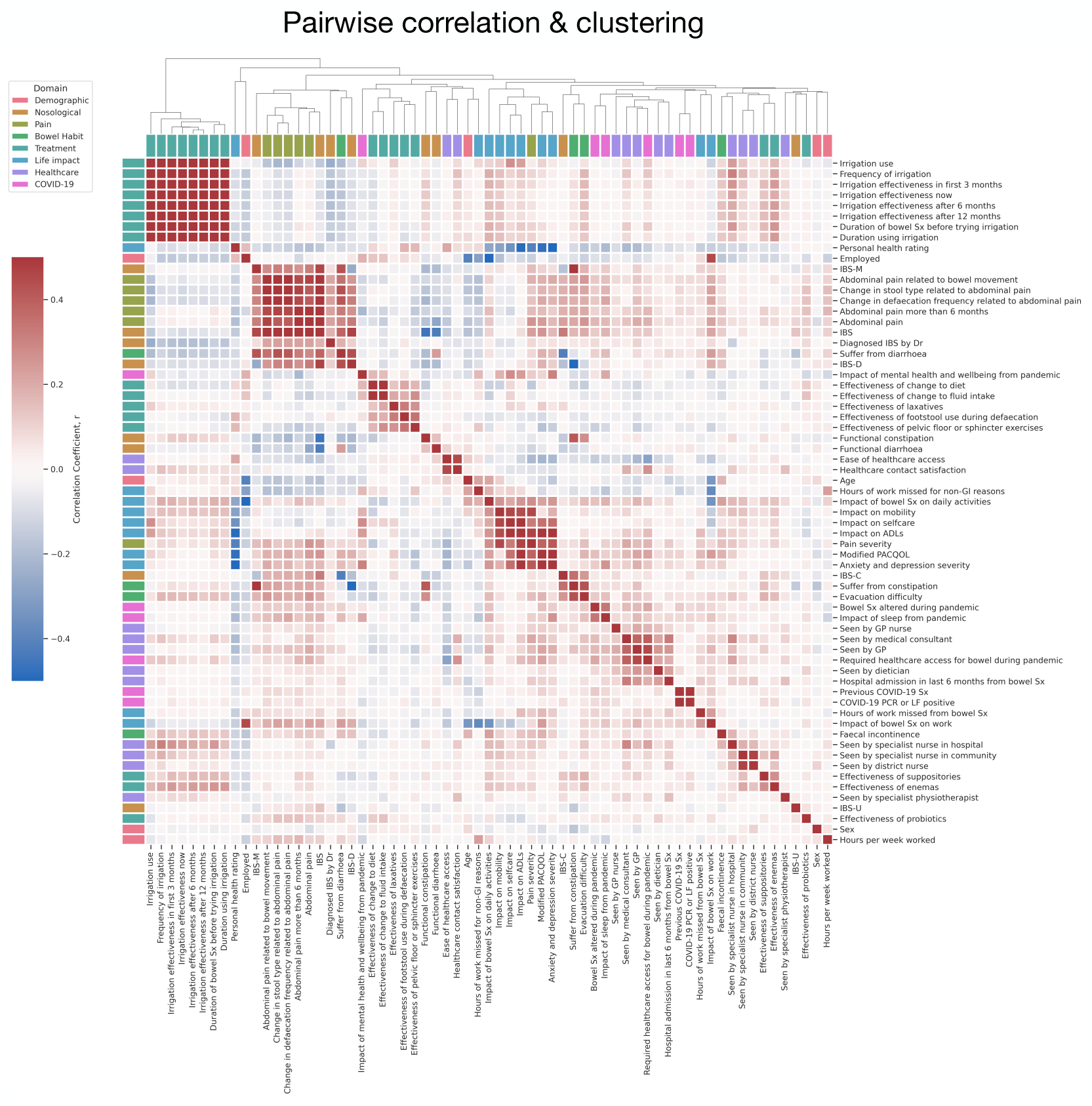
Feature correlation matrix dendrogram. The correlation matrix was derived from the Pearson correlation coefficient, and the hierarchical clustering dendrogram was derived from the Euclidean distance matrix. Darker red squares depict more positive, and darker blue squares depict more negative correlation coefficients between pairwise factors.

Taken together, these initial analyses show that many aspects of patient data cluster together into relatively self-explanatory domains when modelled in pairwise linear terms. Measures of abdominal pain (and FBD diagnoses made by the presence of pain^3^) cluster together. Where an aspect of a patient’s daily life is disrupted, disruption to other aspects of their life is also likely. Where a response to one treatment is identified, there is likely to be some response to another. The key here, however, is that such an approach only superficially characterises pairwise and linear relationships between a patient or disease factor; a remit nonlinear machine models allow us to interrogate further.

### Machine model predictions of all patient factors

The out-of-sample test set performance breakdowns for all models across the different data domains are shown compactly in Figure 3, Table 1, and Table 2 and described in greater detail within the supplementary material. Regression models (Figure 3A, Table 1) tasked to predict healthcare usage and life impact achieved the best out-of-sample predictive performances, with more variable performances across treatment, pain, demographic, and COVID-19 impact domains. These findings illustrate how the healthcare requirements of a given individual could be relatively well determined with machine learning, plausibly applicable to triage systems or healthcare system planning, and similarly how the impact on daily life was relatively easily determined also, of relevance to determining the broader impact of disease at both the individual and societal level, whereas predicting individual treatment response in this cohort was a far more challenging task.

**Figure 3.**
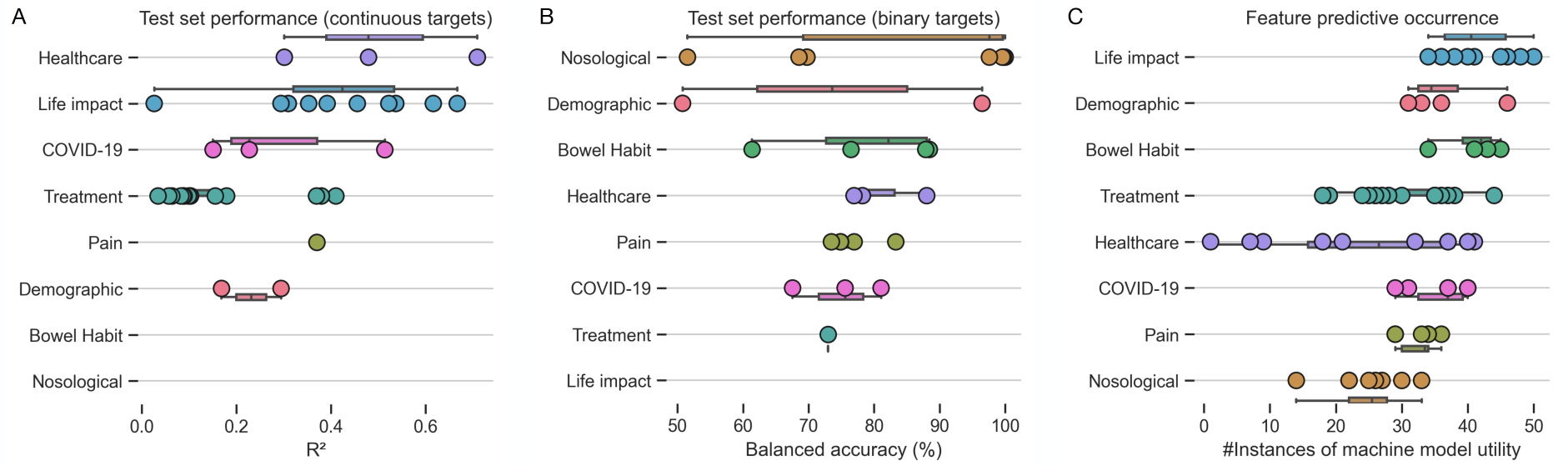
Machine model performances and feature importance across all domains. A) Test-set performance for regression models (in R^2^). B) Test-set performance for classification models (in % balanced accuracy). C) Feature occurrence across all modelling tasks, illustrating the frequent use of life impact measures in machine model predictions, where diagnostic (nosological) data use was uncommon. All panels are stratified and colour-coded by data domain, as shown on the y-axis.

**Table 1.** Out-of-sample test set performances for regression models. Model performance is given by the R^2^ value, where a higher value indicates greater predictability from the remaining patient data. Abbreviations: ADLs, activities of daily living; PACQOL, patient assessment of constipation-quality of life.

| Regression model target | Test set performance (R<sup>2</sup>) |
| --- | --- |
| Frequency of attendance for bowel symptoms | 0.71 |
| Impact of bowel symptoms on daily activities | 0.67 |
| Impact of bowel symptoms on work | 0.62 |
| Anxiety and depression severity | 0.54 |
| Impact on ADLs | 0.52 |
| Impact of mental health and wellbeing from the pandemic | 0.51 |
| Ease of healthcare access | 0.48 |
| Modified PACQOL | 0.46 |
| Effectiveness of pelvic floor or sphincter exercises | 0.41 |
| Impact on selfcare | 0.39 |
| Effectiveness of fluid intake | 0.38 |
| Pain severity | 0.37 |
| Effectiveness of change to diet | 0.37 |
| Personal health rating | 0.35 |
| Hours of work missed for non-GI reasons | 0.31 |
| Healthcare contact satisfaction | 0.30 |
| Age | 0.29 |
| Impact on mobility | 0.29 |
| Impact of sleep from the pandemic | 0.23 |
| Effectiveness of footstool use during defecation | 0.18 |
| Hours per week worked | 0.17 |
| Effectiveness of laxatives | 0.16 |
| Bowel symptoms during the pandemic | 0.15 |
| Effectiveness of probiotics | 0.10 |
| Effectiveness of enemas | 0.09 |
| Effectiveness of suppositories | 0.08 |

**Table 2.** Out-of-sample test set performances for classification models. Model performance is given by percentage balanced accuracy, where a higher value indicates greater predictability from the remaining patient data.

| Classification model target | Test set performance (balanced accuracy %) |
| --- | --- |
| Diagnosis – IBS, and if IBS-D, IBS-M, IBS-C | 100% |
| Employment status | 96% |
| If suffering from diarrhea or constipation | 88% |
| If seen by a GP for FBD | 88% |
| If suffering from abdominal pain | 83% |
| If required healthcare access for bowel reasons during the pandemic | 81% |
| If seen by a specialist hospital nurse | 78% |
| Presence of abdominal pain for six months or more | 77% |
| If seen by a medical consultant for FBD | 77% |
| If suffering with evacuation difficulty | 76% |
| Change in stool types related to abdominal pain | 75% |
| Abdominal pain related to bowel movements | 73% |
| Irrigation use | 73% |
| If diagnosed as IBS by a doctor specifically | 70% |
| Diagnosis - functional constipation | 69% |
| Diagnosis - fecal incontinence | 61% |
| Diagnosis - functional diarrhea | 52% |
| Sex | 51% |

The classification models in panel Figure 3B and Table 2 demonstrate that disease classification (nosology) was overall most predictable, followed by bowel habit, healthcare usage, pain, COVID-19 impact, and treatment data. The high classification accuracy of patient diagnosis is an expected finding, given the clear constellation of signs and symptoms that algorithmically determine them^4^. Yet, the ability to accurately determine employment status, healthcare usage (and type of) at the individual level holds plausible value for quantifying the wider impact the FBDs cause, pertinent to the patient lived experience.

A key advantage of machine learning is its ability to undertake feature selection, automatically choosing the greatest determinants of a given modelling target to build the best-performing model. In reviewing the feature importance and contributions across all model targets, it transpired that whilst patient factors of life impact, demographics, and bowel habit were commonly selected by XGBoost runs, rarely was the patient’s diagnostic label selected, suggesting that diagnosis was, in fact, minimally helpful in predicting wider patient factors, including those of their lived experience (Figure 3C).

### Determinants of symptom burden and quality of life

Models predicted symptom burden and quality of life measures relatively well, relying predominantly on life impact, mental well-being, and age to determine them, but notably, not diagnostic labels. We provide a breakdown of performant models in determining symptom burden and quality of life metrics with SHAP plots in Figure 4, further discussed in the supplementary material.

**Figure 4.**
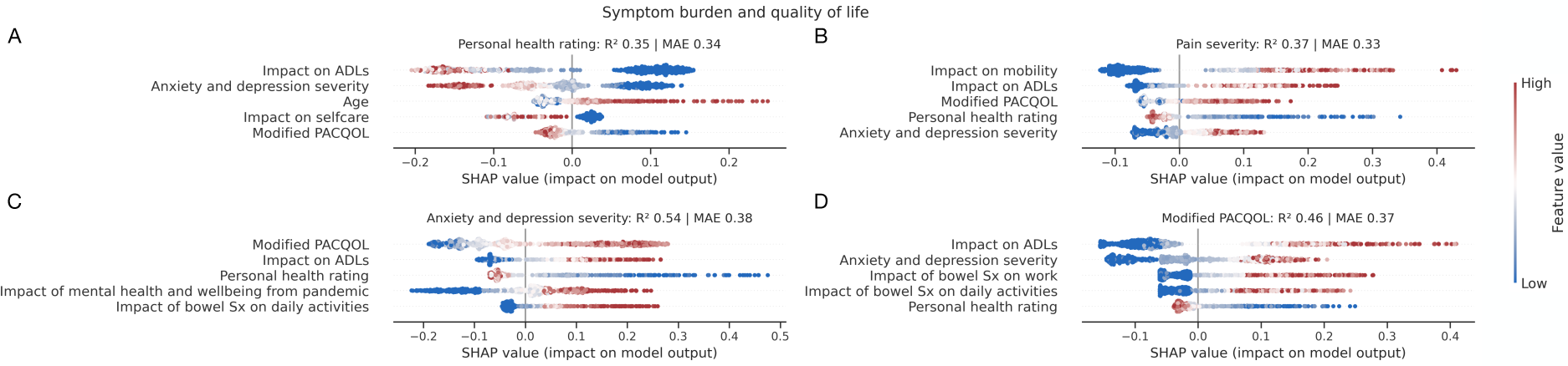
Determinants of symptom burden and quality of life. SHAP plots for machine learning models quantifying patient A) personal health rating, B) pain severity, C) anxiety and depression severity, and D) modified PACQOL. Out-of-sample performance is shown by R^2^ and mean absolute error (MAE). Only the top 5 predictive factors of each target are shown for visualisation purposes. For each panel, each point represents a patient, and each row is an input feature to the model, where positive x-axis values depict a *positive* impact on the model output, and redder points depict higher feature values. For example, panel A) shows the top predictive feature for personal health rating to impact on ADLs, where a greater (i.e., more detrimental) impact on ADLs was associated with the patient reporting a lower (i.e., worse) personal health rating. Key here is that patient diagnosis was not selected by the models as informative in their prediction.

### Determinants of life impact from functional bowel disorders

Models accurately predicted life impact targets, primarily determined by hospital attendance data, employment status, other life impact measures, mental well-being, and pain data, but not diagnostic labels. In Figure 5, we provide a breakdown of performant models in determining life impact with SHAP plots, which is further discussed in the supplementary material.

**Figure 5.**
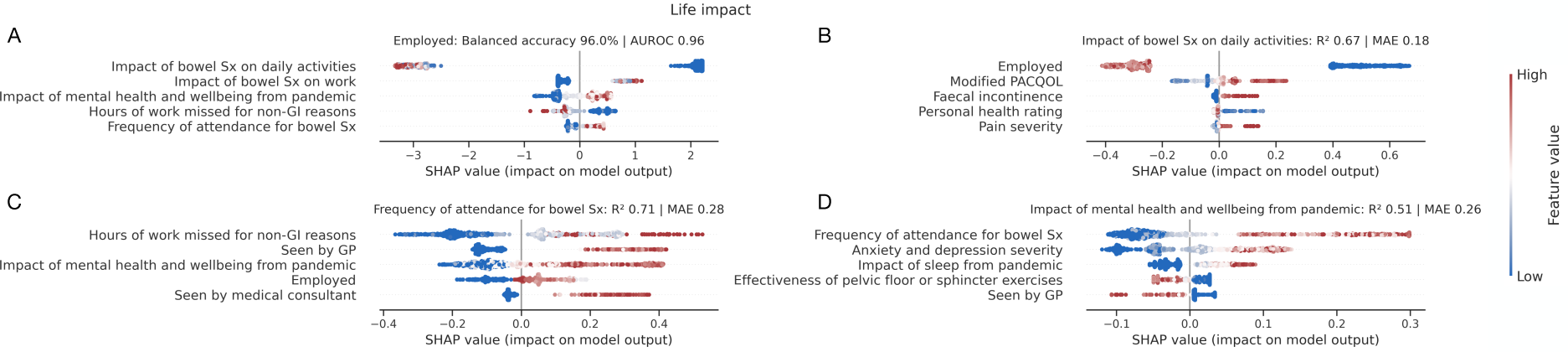
Determinants of life impact. SHAP plots for machine learning models quantifying patient A) employment status, B) impact of bowel symptoms on daily activities, C) frequency of healthcare attendance for bowel symptoms, and D) impact of mental health and wellbeing from the pandemic. Out-of-sample performance is shown by % balanced accuracy and AUROC for classification models (A), with R^2^ and mean absolute error (MAE) for regression models (B-D). Only the top 5 predictive factors of each target are shown for visualisation purposes. A description of interpreting SHAP plots is given in the legend in Figure 4. The patient diagnosis was not selected by the models as informative in their prediction.

### Determinants of patient-reported treatment effectiveness

Model performance in predicting patient-perceived treatment effectiveness was more variable. Despite variable performance, patient-reported treatment response to one intervention was largely predictive of response to another. Conversely, those refractory to one intervention were likely to be refractory to others. In Figure 6, we provide a breakdown of performant models in determining patient-perceived treatment effectiveness with SHAP plots.

**Figure 6.**
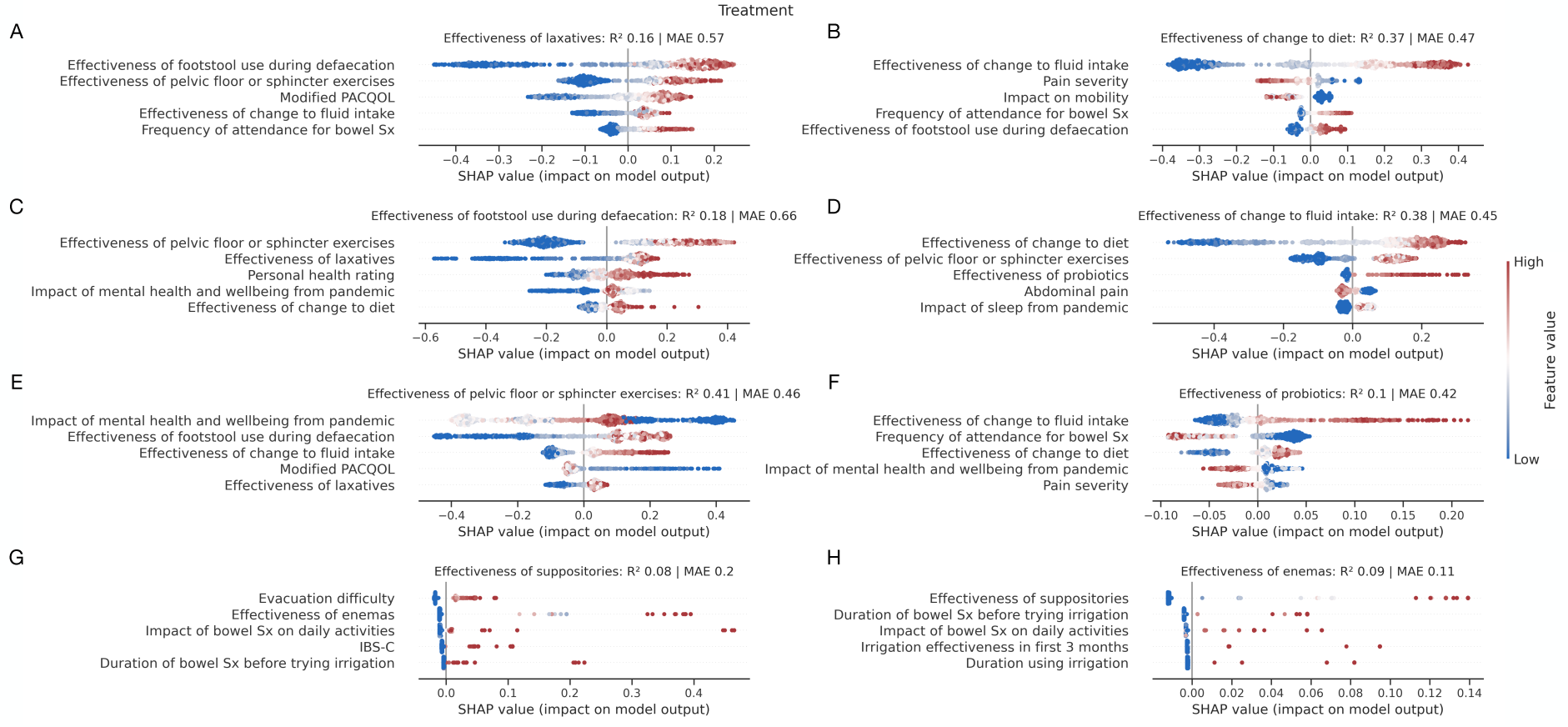
Determinants of treatment response. SHAP plots for machine learning models predicting the effectiveness of A) laxatives, B) dietary changes, C) footstool usage during defecation, D) fluid intake changes, E) pelvic floor or sphincter exercises, F) probiotics, G) suppositories, and H) enemas. Out-of-sample performance is shown by R^2^ and mean absolute error (MAE). Only the top 5 predictive factors of each target are shown for visualisation purposes. A description of interpreting SHAP plots is given in the legend in Figure 4. Diagnosis only features in one of eight treatment models, where predictive performance was also notably poor (panel G).

The foregoing analyses illuminate relationships between singular aspects of the patient’s lived experience. While disclosing non-linear and higher-order relationships between sets of factors in predicting another, the approach lacks an all-encompassing compact summary of the patient’s lived experience that only an unsupervised approach could plausibly offer. Given the interweaving of disease and patient features, this may be best approximated by a generative model of a network.

We therefore fitted a nested generative stochastic block model comprising all factors as nodes, with weighted directed edges as feature contributions to each machine model, revealing a sophisticated community structure of patient factors (Figure 7). This was broadly organised into the domains of nosology, life impact, treatment effects, and symptomology. The network structure reiterated the importance of symptom and life-impact factors instead of diagnosis-related ones.

**Figure 7.**
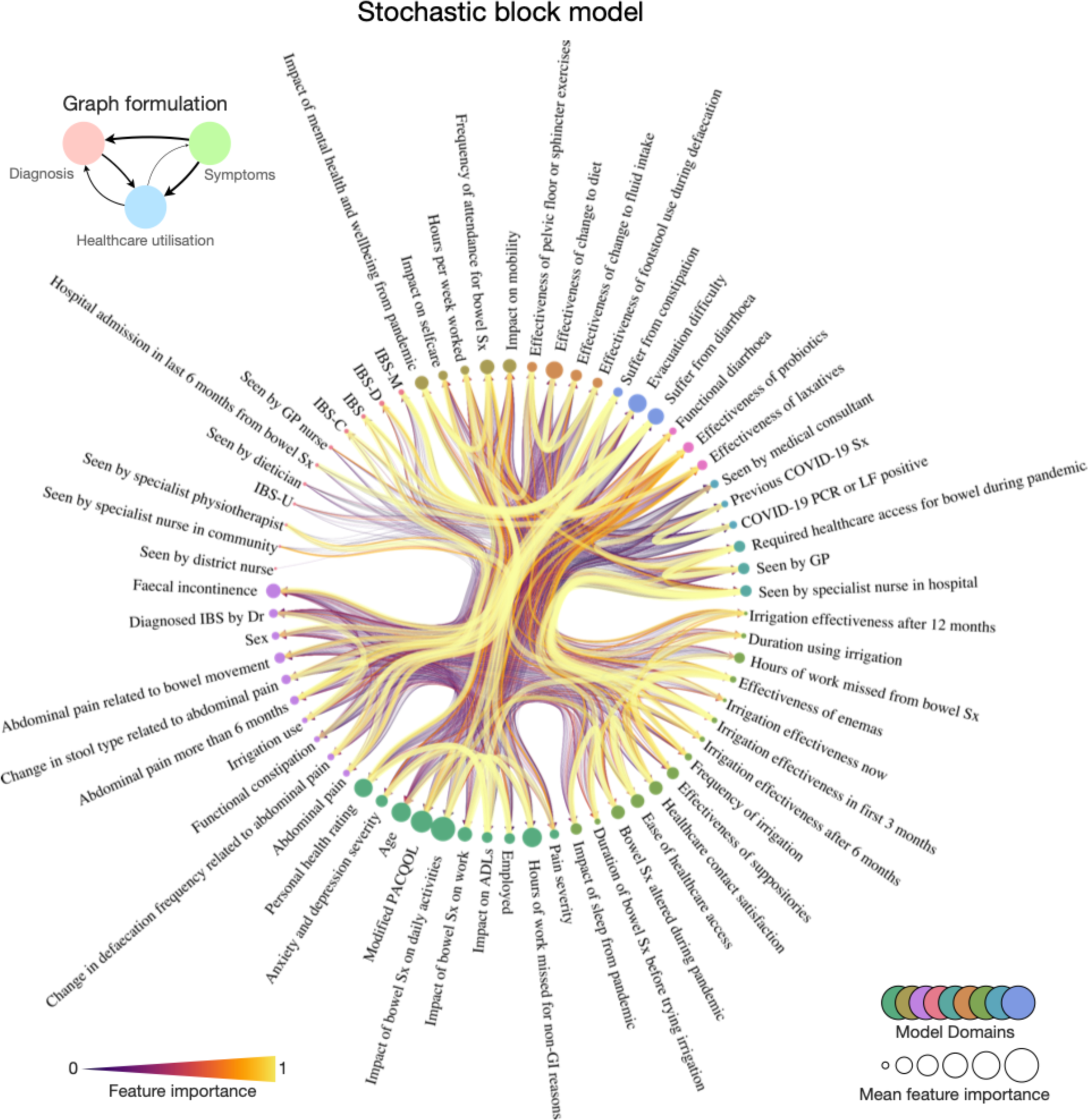
A generative network community structure for the lived experience of functional bowel disorders. Radial network of the nested, generative Bayesian stochastic block model community structure of patient factors. Nodes are individual circles with corresponding text labels, sized according to their importance in predicting all other target nodes. Edges are weighted by the directional feature importance in predicting one feature over another, where edge width and colour are proportional to the key. Node communities are similarly colour-coded as per the key at the second hierarchical level. Supplementary Figure 2 accompanies this figure with additional results.

Next, we extracted each community block’s weighted eigenvector, hub, authority, and centrality metrics at the second nested (L_1_) level. Eigenvector centrality measures a node’s ‘influence’ across the whole network^17^. The Hyperlink-Induced Topic Search (HITS) is a centrality algorithm historically developed for rating worldwide web pages, stemming from the observation that when the internet was initially forming, specific web pages operated as large directories – hubs – yet were not authoritative to the information contained within them, although were indeed helpful as catalogues to direct people to the authoritative pages^18,19^. Framed differently, a ‘good hub’ of a network of the internet points to many other pages, whilst a ‘good authority’ would be a page linked by many different hubs^20^.

This quantitative analysis of our generative network structure found the node community consisting of constipation or diarrheal disease nosology had significantly greater hub centrality than all other node blocks (the measure of how often a node links to other factors irrespective of how informative or authoritative it may be) (one-way ANOVA with post-hoc Tukey p<0.0001) (Figure 8). Meanwhile, two communities consisting of treatment effects and life impact measures had significantly greater eigenvector centrality (the measure of a node’s ‘influence’ across the whole network) (one-way ANOVA with post-hoc Tukey p<0.0001). Similarly, the node community of treatment effectiveness related to probiotics and laxatives and node community related to life impact both yielded significantly greater authority centrality (the measure of how informative and authoritative a node is to the remaining network) (one-way ANOVA with post-hoc Tukey p<0.0001).

**Figure 8.**
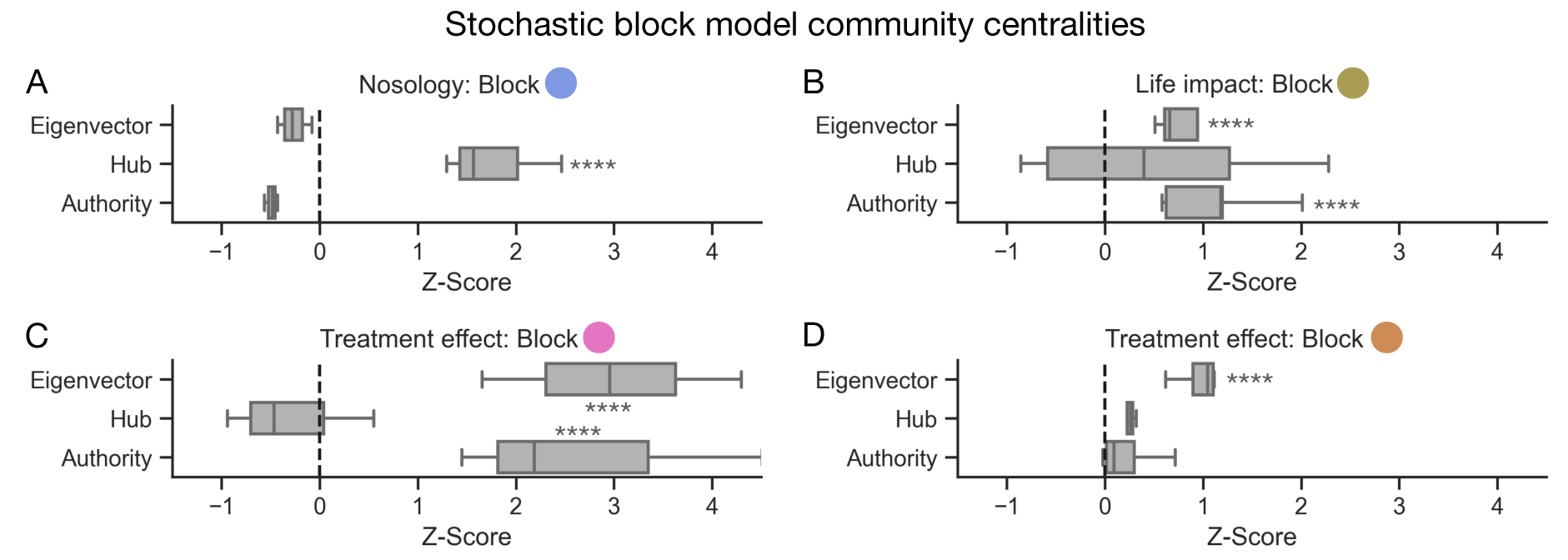
Diagnoses are hubs, but life impacts and treatment effects are authorities and influencers. Box and whisker plots illustrating centrality metrics of A) the nosology node community from the nested stochastic block model comprising if the patient suffers from diarrhoea, constipation, or any form of evacuatory difficulty; B) life-impact including self-care, mobility, ability to work, healthcare access and mental wellbeing; C) the treatment effectivity node community comprising the effects of probiotics, effects of laxatives, and also a diagnosis of functional diarrhoea; and D) the treatment effectivity node community comprising the effects of pelvic floor/sphincter exercises, dietary or fluid changes, and footstool use during defecation. Refer to Figure 7 for a radial representation of this network community structure. **** denotes a post-hoc Tukey significance test of p<0.0001 following one-way ANOVA.

## Discussion

### Deep phenotyping the lived experience, applied to functional bowel disorders

Modern medicine has become an algorithmic science: the clinician assesses symptoms, orders tests, and classifies the illness into a diagnostic category on which therapy is based. In FBDs, there are no objective measures that sub-classify patients based on pathophysiology; instead, symptom-based classifications are crafted to aid management. Whilst this can be helpful, it may over-simplify the portrayal of a complex multi-dimensional condition, where linkage to lived experience is vital^1,2^. We have conducted a holistic characterisation of a large cohort of patients with FBD using a multi-dimensional machine learning approach without prior assumptions on the determinants of the lived experience. The main pertinent findings are twofold:

Firstly, we reveal in unprecedented detail the determinants of patient-reported symptom burden, quality of life, life impact, and treatment effectiveness and provide the framework for predicting them. In many ways, these determinants are at odds with what we, as healthcare professionals, often assign as the determinants of health and wellbeing; examples are discussed below.

Secondly, our generative network analysis, summarising the output of a comprehensive machine modelling framework, reveals the high-dimensional community structure of these patient factors. This process formally quantifies that whilst the nosological domains of disease we classify patients with are network hubs that link many aspects of patient health and wellbeing, they are poorly influential (or authoritative) in describing the broader aspects of patient health. Instead, a focus on patient-reported treatment effectiveness and impact on a patient’s daily life is instead quantitatively authoritative and influential.

The value in developing a suite of machine models to predict these characteristics is not merely the depiction of those with high performance (which is most often the case in machine learning - and digital medicine - research). Rather, this process illuminates what can (but equally importantly, what cannot) be determined from data routinely available in a clinical setting. It should come as no surprise that a diagnosis of IBS can be predicted perfectly from metrics of abdominal pain and gastrointestinal disturbance: these factors are definitional for a diagnosis of IBS by current classification systems^4,6^. More important, however, is the fidelity of models to ascertain other aspects of patient health, such as the effect of young age or employment status, which are not as intuitive.

### The determinants of patient health and well-being

Machine learning models are often described as sophisticated in their ability to formulate a decision based on nonlinear interactions amongst complex multivariate data^13,21^, but the reality is that the healthcare professional reviewing a patient undertakes similar processes to inform their clinical decision-making^13^. To that end—and with a priority of better digital health for these individuals—we have described the machine determinants of ill health from the individual patient’s perspective so that these factors can be considered in the clinical consultation.

#### Symptom burden and quality of life

We find that a patient’s rating of their health is not principally determined by the severity of gastrointestinal symptoms or diagnosis but instead by the impact of disease on their daily life, the presence of anxiety and depression, and being ill at a younger age. While most of these factors are intuitive and unsurprising, the effect of young age on the perception of well-being has not been previously highlighted.

The greatest determinants of patient-reported pain severity were the impact of illness on daily life activities, the presence of anxiety and depression, and the patient’s personal health rating. Quality of life was best determined by the impact of the illness on daily activities, the presence of anxiety and depression, and the impact of bowel symptoms on work. These results formalize the importance of considering not just the gastrointestinal symptoms of FBDs, but rather the impact they exert on patient life.

Other than in suppository use for if there was a diagnosis of IBS-C (a trivial association), the diagnostic label that we as healthcare professionals assign to these patients as part of ‘best practice’ did not feature as a top 5 determinant for any machine model. Indeed, none of the top five determinants of a patient’s health rating specifically interrogated gastrointestinal symptoms, emphasizing the importance of quantifying the impact of life factors (such as employment and daily life) and mental well-being throughout their routine clinical care.

#### Life impact

Patient employment status could be determined by a machine learning model (the model correctly predicted employment status in 229 of 235 out-of-sample test cases). The greatest determinant of employment status was, by some distance, the degree of impact of bowel symptoms on daily activity. Conversely, the greatest determinant of patient-reported impact of bowel symptoms on daily activities was unemployment. It therefore seems reasonable to suggest that simply ascertaining employment status is an especially strong predictor of life impact measures and something that should be considered in every consultation.

One of the highest-performing machine models was in delineating the frequency of patient healthcare attendance for bowel symptoms (which achieved an out-of-sample R^2^ of 0.71). The greatest determinants of healthcare attendance were the hours of work missed, whether the patient had already been seen by a GP, the impact on their mental health, employment status, and if they had seen a medical consultant before. Vitally, the determinants of patient frequency of healthcare attendance for bowel symptoms were not predicted by bowel symptomology, but instead a combination of healthcare access/gatekeeping (i.e., if already known to the service), impact on employment and life, and mental wellbeing. This is in keeping with the previously known evidence of healthcare-seeking in IBS^22,23^. Naturally, quantifying healthcare requirements is essential, as it has implications in designing, planning, and budgeting healthcare service provisions^24^.

Lastly, we briefly draw focus to our model predicting the impact of mental health and well-being during the COVID-19 pandemic. The strongest determinant in this cohort was the frequency of attendance for bowel symptoms. Put another way, mental health in this FBD population was most significantly affected by the ability to access healthcare during a time when the health service was under particular strain. This finding conveys the priority we should place on widening and maximising patient healthcare access to safeguard mental well-being.

#### Treatment effectiveness

Several models aimed to quantify patient-reported effectiveness of routinely provided FBD treatments, both medicinal (laxatives, enemas, suppositories, probiotics) and non-medicinal (dietary or fluid changes, footstool use during defecation, and pelvic floor/sphincter exercises). We would be wary of drawing conclusions when comparing the effectiveness of certain treatments over others, for the study was not designed as a clinical trial to facilitate this. However, the critical insight was in the determinants of patient-reported treatment effectiveness. Namely, the greatest determinant of patient response to *any* treatment was response to *any other*. Once again, symptomatology and disease classification were minimally predictive of treatment response, but other factors (social, psychological, and comorbid) combined to create, in some patients, a state of refractoriness. It is these patients who respond poorly to all treatments that are seen in secondary and tertiary care and require a holistic approach. More research is needed to understand refractoriness in FBD.

### The interacting structure of patient health and well-being

We harmonised the findings to construct a Bayesian generative network revealing the community structure of factors affecting those living with FBDs. Broadly, these feature communities coalesce to nosology, life impact, treatment effects, and symptomology. Interestingly, however, we show that the nosological branch of these factors – i.e., the domains of disease leading to diagnostic labels we assign to these patients – are ‘hubs’ in this network (a feature that points to many others) but are, in fact, minimally influential^17,18,25^. Instead, the communities of life impact and treatment effectiveness are quantitatively more influential (with higher eigenvector centrality) and authoritative (with higher authority centrality) in the remaining aspects of their health. These findings suggest that the assignment of disease labels to these patients is often minimally helpful in disclosing or influencing the broader FBD lived experience, as shown elsewhere in the comparison of constipation-predominant irritable bowel syndrome and functional constipation^6,26^. Instead, we should focus more on reducing life impacts and improving treatment effects through a holistic approach.

### Study limitations

The study quantified a breadth of patient factors feasibly acquired during routine clinical care, ranging from demographic, diagnostic, symptomology, quality of life, healthcare access, and patient-reported effectiveness of regularly given treatments. One limitation is that we did not quantify more comprehensive and/or specialist investigations (e.g., microbiota, gastrointestinal imaging, genetic, or an exhaustive list of comorbidity data) since this would have limited applicability and generalizability of the findings to centres in which these are not part of routine care. To our strength, in maximising the quantification of variables that *are* routinely available in a real-world healthcare setting, we were able to sample a large cohort from which we could construct a suite of machine learning models with proven fidelity that could be evaluated and/or deployed in similar centres in further research.

Secondly, the study was not designed to clinically trial specific treatments. Instead, it was designed as a cross-sectional study, where patients could self-report the effectiveness of the therapies they had experienced throughout their care. This limits inference that could be drawn from an experimental allocation, i.e., a randomised controlled trial, but it does quantify the response to treatment with explicit emphasis on the individual patient experience (as opposed to any biochemical/investigatory endpoint). In any case, our focus was to illuminate the determinants of patient-reported response to treatment *in general* instead of quantifying the superiority/non-inferiority of one treatment over another.

Thirdly, our study was not designed to investigate directional effects. One might suggest a plausible directionality of impaired GI health leading to a triad of increased healthcare utilisation, loss of productivity/employment, and worsening QoL/mental well-being, but it must conform to the appropriate criteria for establishing causality^27^. An additional analytical route would be dedicated causal inference, a task for future research^28^.

## Conclusion

We develop a machine learning and Bayesian generative network approach for deep phenotyping patient disease and lived experience, first applied to characterise FBDs. Our framework reveals new insights into the determinants of patient-reported symptom burden, quality of life, impact on daily life, and treatment effectiveness in a large representative cohort. Strikingly, these determinants are often at odds with what we, as healthcare professionals, typically pre-suppose are the greatest determinants of patient-reported health. Instead of disease classification or symptom severity, this was defined by the impact on daily life, employment status, access to healthcare, and mental well-being. To safeguard mental well-being, patient access to healthcare must be attainable, and a holistic approach to the consultation is required. Patients tend to be responsive to multiple therapies or refractory to all, and a deeper understanding of refractoriness should form a future research priority. In making our approach openly available, we hope to catalyse further study of the patient’s lived experience across broader health remits.

## Methods

### Study design

A single online questionnaire was administered to two existing cohorts of individuals using convenience sampling methods. The two groups were:

1. ContactMe-IBS (established 2017) – a national irritable bowel syndrome (IBS) registry of people who are interested in participating in IBS research (https://www.contactme-ibs.co.uk). ContactMe-IBS is owned by the NHS (County Durham and Darlington NHS Trust). Registrants are primarily from the Northeast (actively promoted within Durham Bowel Dysfunction Service) and the Southwest, where GPs are particularly research active with ContactMe-IBS. Access to the registry is available via numerous sources, including GP practices, gastroenterology clinics, pharmacies, and social media. During registration, participants self-identify as having IBS by completing screening questions based on Rome IV criteria^4^.
2. Transanal irrigation (TAI) database (established 2019) – a database of patients who have commenced TAI under the care of Durham Bowel Dysfunction Service.

Participants on the registry received primary or secondary care for IBS and gave permission to be informed of active research studies. Over four weeks, October - November 2021, registrants of both databases (n = 4480 on ContactMe-IBS; n = 259 on the TAI database) were invited to participate by email link to a questionnaire or postal questionnaire if preferred. Online questionnaire data were captured digitally via the web-based REDCap application, a secure system designed to support data collection for research studies. Inclusion in the study required participants to be 18 years or older with symptoms of bowel dysfunction, registered on either database and able to understand written and spoken English (for questionnaire completion). Participants who did not respond to the invitation or reminder email or those who did not fully complete the questionnaire were excluded.

### Materials

The study used an 88-item questionnaire requiring ∼35 minutes to complete, organised in the following sections:

1. *Demographic*: including date of birth, sex, ethnicity, and employment status.
2. *Nosological*: this section was designed to characterise the FBD type of the participant. The scoring algorithms of the ROME IV^4^ criteria were used to identify primary diagnostic groups: irritable bowel syndrome (constipation [IBS-C], diarrhoea [IBS-D] predominant, or mixed [IBS-M]); functional constipation (FC); functional diarrhoea (FD); or faecal incontinence (FI). Criteria for evacuatory dysfunction (ED) did not depend on investigations but relied on symptom scores for straining, a feeling of blockage, a feeling of incomplete evacuation and the need to digitate, with questions and scoring of these aligned to the ROME IV questionnaire.
3. *Primary symptom:* respondents were asked to report their primary symptom from a choice of ‘abdominal pain’, ‘bloating’, ‘watery stools’, ‘hard stools’, and ‘frequent bowel movements’, including quantified severity and duration experienced.
4. *Bowel habit*: the Bristol Stool Form Scale was used to identify stool type^29^. The ROME IV criteria individual question data was used to assess bowel habit^4^.
5. *Treatment*: A visual analogue scale (VAS) was used to measure perceived effectiveness for a range of trialled treatments. These included medicinal (such as laxative, enema, suppository) and non-medicinal (such as pelvic floor/sphincter exercises, footstool use during defecation, fluid and/or dietary changes). The study team developed questions on the use and effectiveness of TAI for managing FBDs, consisting of seven single-answer multiple-choice questions and a VAS for patient-perceived effectiveness. Data were not curated or designed for treatment comparisons but rather to delineate the determinants of patient-perceived effectiveness to a given regime.
6. *Life impact*: the impact of FBDs on QoL was assessed using a 5-point Likert scale based on the Patient Assessment of Constipation on Quality of Life (PAC-QOL) questionnaire^8,30^. PAC-QOL wording was widened to reflect all FBDs; for example, ‘constipation’ was amended to ‘bowel symptoms’, and questions related directly to constipation were omitted (Q2, Q4, Q20, Q21, Q24 from PAC-QOL^30^). This approach would enable insight into the impact of any set of bowel symptoms on a patient rather than placing focus on specific disease subtypes. The EQ5D-5L General Health^31^ was used to explore the impact of FBDs (on mobility, self-care, usual activities, pain or discomfort, and anxiety or depression. Each patient’s rating of their overall health was also measured by VAS. The Work Productivity and Activity Impairment Questionnaire^32^ assessed impairment in activities of daily living (ADL) and employment-related productivity. Questions elicited employment status, absenteeism (percentage of work hours missed due to bowel symptoms), presenteeism (the degree to which symptoms affect work productivity whilst working), percentage of work hours missed for other reasons, and the degree to which symptoms affected other ADLs in the preceding seven days.
7. *Healthcare use*: questions determined whether the participant had been admitted to hospital for bowel symptoms; and their access to healthcare including physiotherapy, general practitioner (GP), consultant gastroenterologist, GP/district/specialist nurse, and dietician.
8. *COVID-19*: comprising single response multiple choice questions explored how the COVID-19 pandemic affected individuals.

### Algorithmic approach

FBDs are a complex set of disorders that are both impacted by and have a profound impact on a wide array of interacting biological, social, and psychological factors. A study seeking to model or characterise *one* single constitutional, diagnosis, disease, treatment, life impact, or healthcare access feature in such a cohort could only increase its understanding by small margins. Our task here is to find a means to understand these patients’ disease processes and lived experiences in a much broader sense, developing a suite of statistical models aiming to predict *all* patient factors instead.

In undertaking such an approach, we forgo any clinical assumptions, instead harnessing a data-driven method that allows machine models to identify which constitutional, diagnostic, disease, treatment, life impact, or healthcare access factors are predictable whilst simultaneously revealing their data-driven determinants. Our framework tests the hypotheses that 1) a machine model shall discern what patient factors plausibly can – and perhaps equally important, what cannot be – predicted from their remaining data, and 2) a machine model shall identify the greatest determinants of a given patient feature, both of which have downstream clinical utility in decision support, patient monitoring, and treatment.

A practical example is the prediction of patient-reported health quality. In determining the extent to which patient-reported health quality can be predicted from other constitutional and clinical data, this reveals the capacity for healthcare professionals to ascertain it from data routinely available. Where patient-reported health quality is readily predictable by a machine model, then its determining factor(s) can guide practice, whether for patient triaging or treatment monitoring. If, however, health quality is not predictable, then this informs us that data currently curated, inclusive of the patient constitution, diagnosis, and healthcare access, do not inform it, so in our practice, we should not make assumptions as to how a patient would rate their personal health without seeking further information.

We therefore developed a software-embodied, end-to-end, multivariate pipeline to fully interrogate patient data, inclusive of data organisation and multivariate missingness imputation, yielding a platform of machine learning extreme gradient boosting (XGBoost) models optimised to predict a set of given inputs, all evaluated out-of-sample on a separate test set (Figure 1)^33^. XGBoost is an architecture that employs a parallel ensemble of gradient-boosted weak-learner decision trees, which has shown superior performance in multiple machine learning tasks with tabular data^33^. We partitioned data 80:20 into model training (n=940) and testing (n=235) sets; the latter was completely excluded through all model development and evaluated only after completing the development of all models. This pipeline is described in significantly greater detail within the supplementary material.

Our algorithmic approach yielded a comprehensive set of machine models trained to predict each patient feature from the remainder, importance metrics (SHapley Additive exPlanations (SHAP)^34^) which shed light on the strongest determinants of each feature and model performance metrics, all evaluated out of sample. We then consolidated these findings formally with a nested stochastic block model (SBM), a Bayesian generative model of a network that aims to find the most optimum community structure^35-41^. Just as the London Underground network comprises stations (nodes) and tracks between them (edges), organised by different train lines, we study patient factors as nodes and the prediction importance metrics as edges connecting factors to one another. Fitting an SBM to these data reveals the most compact representation of how patient factors are organised, grounded in information theory and Ockham’s razor^37^. We further quantify graph centrality metrics aligned to each community partition to study these organisations of disease and lived experience factors even deeper.

## Data and code availability

All code will be made publicly available upon publication at https://github.com/jamesruffle/perspective-ai. Trained model weights are available upon request. Data and code availability align with UK government policy on open-source code. Patient data are not available for dissemination under the ethical framework that governs its use.

## Ethical approval

The study was approved by the local institutional review board and conducted in accordance with the Declaration of Helsinki. The Health Research Authority approved this study prior to commencement. REC reference 21/SW/0086 (IRAS ID 296856).

## Manuscript Type

Original article

## Data Availability

All code will be made publicly available upon publication at https://github.com/jamesruffle/perspective-ai. Trained model weights are available upon request. Data and code availability is in line with UK government policy on open-source code. Patient data are not available for dissemination under the ethical framework that governs its use.

## Supplementary Material

### Method

#### Algorithmic approach

##### Handling of data missingness

In line with standard practices, we removed patients with missing data for >40% of feature columns (n=1) and conversely removed features with missing data for >40% of patients (f=19), resulting in 59 features. We established that the data was missing at random (MAR) and used multivariate imputation via chained equations (MICE)^1^ with predictive mean matching, 50 multiple imputations and 50 iterations (i.e., x10 the default for each) to impute the remaining missing values.

##### Visualisation of the patient-feature space

We next created a visualisation of the feature space across all patient domains of demographic, nosological, pain, bowel habit, treatment, life impact, healthcare, and COVID-19 in the form of a cluster map. We computed the pairwise Pearson correlation matrix of all features and performed hierarchical clustering by Euclidean distance metrics to produce a dendrogram^2-5^. P values were adjusted by False Discovery Rate^6^. For cross-group comparisons, including patient-reported treatment effectivity, we conducted one-way ANOVAs with post-hoc Tukey testing.

##### Machine learning

While Euclidean distance-derived dendrograms can display linear, pairwise feature relationships, we anticipated that FBD complexity would necessitate machine learning models capable of capturing non-linear, interacting relationships within the feature space.

##### Data pre-processing

We partitioned data into 80:20 train (n=940) and test (n=235) sets; the latter was completely held out and evaluated only after the complete development of all models. All hyperparameter optimisation was performed using cross-validation in the training set only.

We removed features directly related to the target with regular expression to avoid trivially reductive predictions. For example, if the target involved irrigation, all predictive features involving irrigation were dropped. We retained features explicitly related to a given diagnosis (e.g., the presence of abdominal pain when predicting IBS) to be used as benchmark models, ensuring appropriate fidelity with the modelling architectures and hyperparameters^7,8^.

Targets where the class imbalance exceeded 20:1 were excluded entirely due to insufficient data support. For those remaining, we used Synthetic Minority Over-sampling (SMOTE) to handle class imbalances^9^. This well-established technique oversamples the minority class by creating new cases over a learnt linear manifold^9^. All data were clamped along the 0.1th and 99.9th percentile and normalised to limit the influence of extreme outliers and scale variance, in line with standard practices.

##### Model training

For all predictive modelling, we used eXtreme Gradient Boosting (XGBoost)^10^, an architecture that employs a parallel ensemble of gradient-boosted weak-learner decision trees. This architecture has shown superior performance in multiple machine learning tasks^10^. Models were constructed as classifiers for categorical targets and regressors for continuous targets. All training runs of a single XGBoost model (for one given target, e.g., patient sex) were undertaken with 5-fold cross-validation. Hyperparameters were optimised by gridsearch across learning rate, number of estimators, maximum depth, subsampling, gamma, and the minimum sum of the instance weight (inside the 80% training data partition), using negative log loss for classifiers and root mean squared error for regressors. This required 16,200 individual model fits per target, which across all 59 targets equated to 955,800 model fits, which took approximately four days on a NVIDIA 2080Ti GPU.

##### Feature importance

We derived feature importance scores for all model runs by the number of times a feature is used to split the data across all trees (i.e., the XGBoost default^10^) and feature directionality with SHapley Additive exPlanations (SHAP)^11^.

##### Out-of-sample evaluation

After identifying the optimum fit, we evaluated model performance on the test set for classifiers deriving the balanced accuracy, precision, recall, and F1 (all macro-averaged) and for regressors deriving the mean absolute error, mean squared error, RMSE, and R^2^. MAE and MSE were reported as a function of normalised and z-scored targets to facilitate performance comparison across targets with varying ranges (e.g., the age range was 20-85 years, whereas personal health rating was between 6-100).

##### Bayesian generative graph models of complex nonlinear feature relationships

The preceding analyses characterise feature relationships criterion on predicting a singular task: the gender of a patient, their quality of life, burden of disease, *et cetera*. Whilst innovative in disclosing the non-linear and interacting relationships between sets of features in predicting another, the approach lacks an all-encompassing compact summary that only an unsupervised approach could offer. To that end, we turned to graph theory as our solution. Graph theory provides a powerful method of modelling complex systems that combines flexibility with intelligibility^12-17^. It treats individual factors of interest as the “nodes” of a network and their interactions as the connections, or “edges”, between them. Here, nodes were all patient features, and edges were the feature importance indices from XGBoost predictive models, the importance metric of one feature in predicting another. The value of this approach over simple metrics of pairwise similarity (e.g., correlation) is that each edge between features captures the importance of that feature in predicting another. This definitionally incorporates the impact of other features in the graph to yield a high-dimensional community structure whilst formally implementing Occam’s razor by Bayesian inference^17^.

We evaluated the community structure of this graph with a non-parametric Bayesian hierarchical weighted stochastic block model. Having derived a community structure of patient features, we extracted block partitions and derived weighted centrality metrics. These centrality metrics were: i) eigenvector (a measure of node ‘*influence’* on the overall graph), ii) authority centrality (a measure of node *authority* in information to other nodes), and iii) hub centrality (a measure of the propensity for a node to link many other nodes). The full mathematical derivation of these metrics is beyond the scope of this article but well established and discussed in significantly further detail elsewhere^2,12,13,18-20^.

A stochastic block model (SBM)^21^ is a generative model of the community structure of a graph composed of *N* nodes, divided into *B* blocks with edges *e*_*rs*_ between blocks *r* and *s*. The model can be framed hierarchically, where edge counts *e*_*rs*_ form block multigraphs with nodes corresponding to individual blocks and edge counts arising as edge multiplicities between block pairs, including self-loops. We seek to infer the most plausible partition (*b*_*i*_) of the nodes, where (*b*_$_) ∈ [1, *B*]^%^ identifies the block membership of node *i* in observed network *G*, with maximisation of the posterior likelihood *P*(*G*|(*b*_$_)). The result is a hierarchically organised community structure of nodes assigned into blocks that yields the most compact representation of the graph, as indexed by its minimum description length^22^, ∑. The general approach is described in further detail elsewhere^21^. Directed feature importance weights were modelled as exponential. Having initialised a fit, we used simulated annealing to optimise it, with a default inverse temperature of 1 to 10. We did not specify a finite number of draws; rather, we specified a wait step of 1000 iterations for a record-breaking event to ensure that equilibration was driven by changes in the entropy criterion instead of driven by a finite number of iterations as per^2,12,23^.

#### Software

Analyses were predominantly performed within a Python (version 3.6.9) environment with the following software packages: graph-tool^24^, imblearn^25^, Matplotlib^26^, NumPy^27^, pandas^28^, SciPy^4,29^, Scikit-learn^29^, seaborn^3^, SHAP^11^ and XGBoost^10^. MICE was performed in R (version 4.1.3)^1^.

#### Hardware

Analyses were predominantly performed on a local 32-core Linux workstation housing 135Gb of RAM and an NVIDIA 2080Ti GPU.

### Results

**Supplementary Figure 1:**
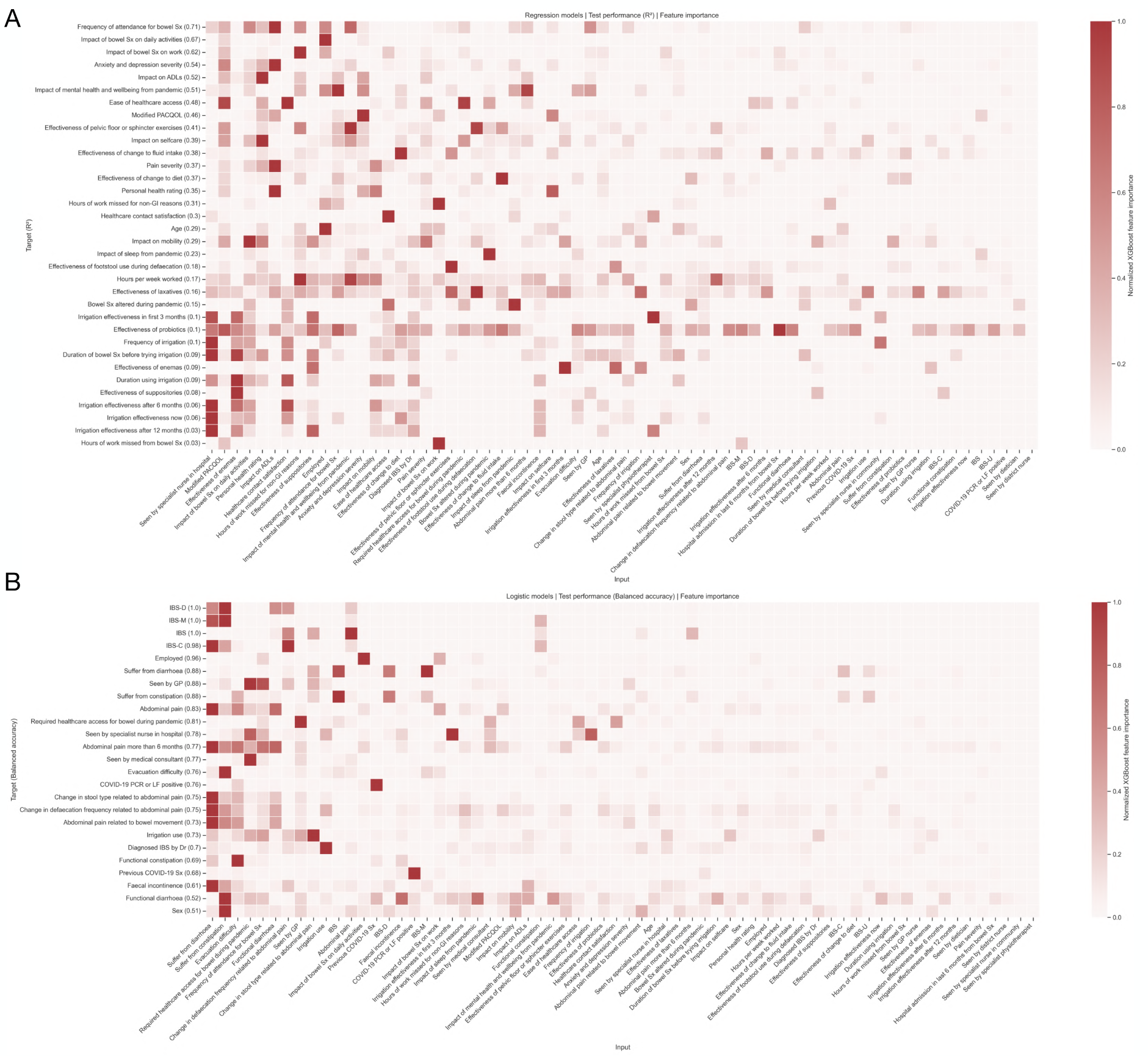
Machine model performances and feature importance heatmaps. Heatmaps of a) regression and b) classification machine models, where the target is shown on the y-axis, and the input feature is shown across the x-axis. In regression models (a), the y-axis target order is per the out-of-sample R^2^ performance, whereas in classification models (b), the order is by out-of-sample balanced accuracy (bracketed values). Input features across the x-axes are coloured by the XGBoost feature importance index, with darker squares more important in target prediction. The X-axis is sorted by mean-feature importance: features with higher mean importance scores in predicting the y-axis target tend to the left side of the heatmap.

#### The determinants of symptom burden and quality of life

First, a machine model could predict a patient’s personal health rating with an out-of-sample performance of R^2^ of 0.35 and MAE of 0.34. The top five determinants of personal health rating were, in descending order: i) impact on ADLs, ii) anxiety and depression severity, iii) age, iv) impact on self-care, and v) modified PACQOL. A machine model could predict pain severity with an out-of-sample performance of R^2^ of 0.37 and MAE of 0.33. The five most predictive determinants of patient-reported pain severity were, in descending order: i) impact on mobility, ii) impact on ADLs, iii) modified PACQOL, iv) personal health rating, and v) anxiety and depression severity. A machine model could also predict anxiety and depression severity with an out-of-sample performance of R^2^ of 0.54 and MAE of 0.38. The five most predictive determinants of anxiety and depression severity were, in descending order: i) modified PACQOL, ii) impact on ADLs, iii) personal health rating, iv) impact of mental health and wellbeing from the pandemic, and v) impact of bowel symptoms on daily activities. Lastly, a machine model could predict PACQOL with an out-of-sample performance of R^2^ of 0.46 and MAE of 0.37. The five most predictive determinants of PACQOL were, in descending order: i) impact on ADLs, ii) anxiety and depression severity, iii) impact of bowel symptoms on work, iv) impact of bowel symptoms on daily activity, and v) personal health rating.

#### The determinants of life impact from functional bowel disorders

A machine model could predict patient employment status with an out-of-sample balanced accuracy of 96% and AUROC 0.96. The five most predictive determinants employment status were, in descending order: i) impact of bowel symptoms on daily activities, ii) impact of bowel symptoms on work, iii) impact of mental health and wellbeing from the pandemic, iv) hours of work missed for non-GI reasons, and v) frequency of attendance for bowel symptoms. A machine model could predict the impact of bowel symptoms on daily activities with an out-of-sample performance of R^2^ of 0.67 and MAE of 0.18. The five most predictive determinants of impact on daily activities from bowel symptoms were, in descending order: i) employment status, ii) modified PACQOL, iii) faecal incontinence, iv) personal health rating, and v) pain severity. A machine learning model could predict the frequency of patient attendance for bowel symptoms with an out-of-sample performance of R^2^ of 0.71 and MAE of 0.28. The five most predictive determinants of attendance were, in descending order: i) hours of work missed for non-GI reasons, ii) if already seen by a GP, iii) impact of mental health and wellbeing during the pandemic, iv) employment status, and v) if previously seen by a medical consultant. Lastly, a machine learning model could predict the impact of patient mental health from the pandemic with an out-of-sample performance of R^2^ of 0.51 and MAE of 0.26. The five most predictive determinants of mental health and wellbeing during the pandemic were, in descending order: i) frequency of attendance for bowel symptoms, ii) anxiety and depression severity, iii) impact of sleep from the pandemic, iv) effectiveness of pelvic floor or sphincter exercises, and v) if seen by a GP.

#### The determinants of treatment effectiveness

Patient reported effectiveness of laxatives could be predicted with an out-of-sample performance of R^2^ of 0.16 and MAE of 0.57. The five most predictive determinants of laxative effectiveness were, in descending order: i) effectiveness of footstool use during defaecation, ii) effectiveness of pelvic floor or sphincter exercises, iii) modified PACQOL, iv) effectiveness of change to fluid intake, and v) frequency of attendance for bowel symptoms. The effectiveness of diet changes could be predicted with an out-of-sample performance of R^2^ of 0.37 and MAE of 0.47. The five most predictive determinants of diet change effectiveness were, in descending order: i) effectiveness of change to fluid intake, ii) pain severity, iii) impact on mobility, iv) frequency of attendance for bowel symptoms and v) effectiveness of footstool use during defaecation. Meanwhile, the effectiveness of footstool use during defecation could be predicted with an out-of-sample performance of R^2^ of 0.18 and MAE of 0.66. The five most predictive determinants of footstool effectiveness were, in descending order: i) effectiveness of pelvic floor or sphincter exercises, ii) laxative effectiveness, iii) personal health rating, iv) impact of mental health and wellbeing during the pandemic, and v) effectiveness of change to diet.

Patient reported effectiveness of changes to fluid intake could be predicted with an out-of-sample performance of R^2^ of 0.38 and MAE of 0.45. The five most predictive determinants of effectiveness in modifying fluid intake were, in descending order: i) effectiveness of change to diet, ii) effectiveness of pelvic floor or sphincter exercises, iii) effectiveness of probiotics, iv) abdominal pain, and v) impact of sleep from the pandemic. Patient reported effectiveness of pelvic floor or sphincter exercises could be predicted with an out-of-sample performance of R^2^ of 0.41 and MAE of 0.46. The five most predictive determinants of effectiveness in pelvic floor or sphincter exercises were, in descending order: i) impact of mental health and wellbeing from the pandemic, ii) effectiveness of footstool use during defaecation, iii) effectiveness of fluid intake changes, iv) modified PACQOL, and v) effectiveness of laxatives. Lastly, the patient-reported effectiveness of probiotics, suppositories, and enemas could only be predicted with an out-of-sample performance of R^2^ of 0.10 and MAE of 0.42; R^2^ of 0.08 and MAE of 0.20; and R^2^ of 0.09 and MAE of 0.11, respectively. The five most predictive determinants of effectiveness in probiotic use were, in descending order: i) effectiveness of change to fluid intake, ii) frequency of attendance for bowel symptoms, iii) effectiveness of change to diet, iv) impact of mental health and wellbeing during the pandemic, and v) pain severity. Relatively few patients reported effects with enemas and suppositories, so we were wary of drawing much inference as to their determinants of positive effect. For completion, we provide these in Figure 6 but suggest some caution in their interpretation under the smaller sampling size.

#### Generative graph community structure

The model entropy (goodness of fit criterion) was -635.54 nats after MCMC by simulated annealing, with an iteration curve indicative of model convergence (Supplementary Figure 2).

**Supplementary Figure 2.**
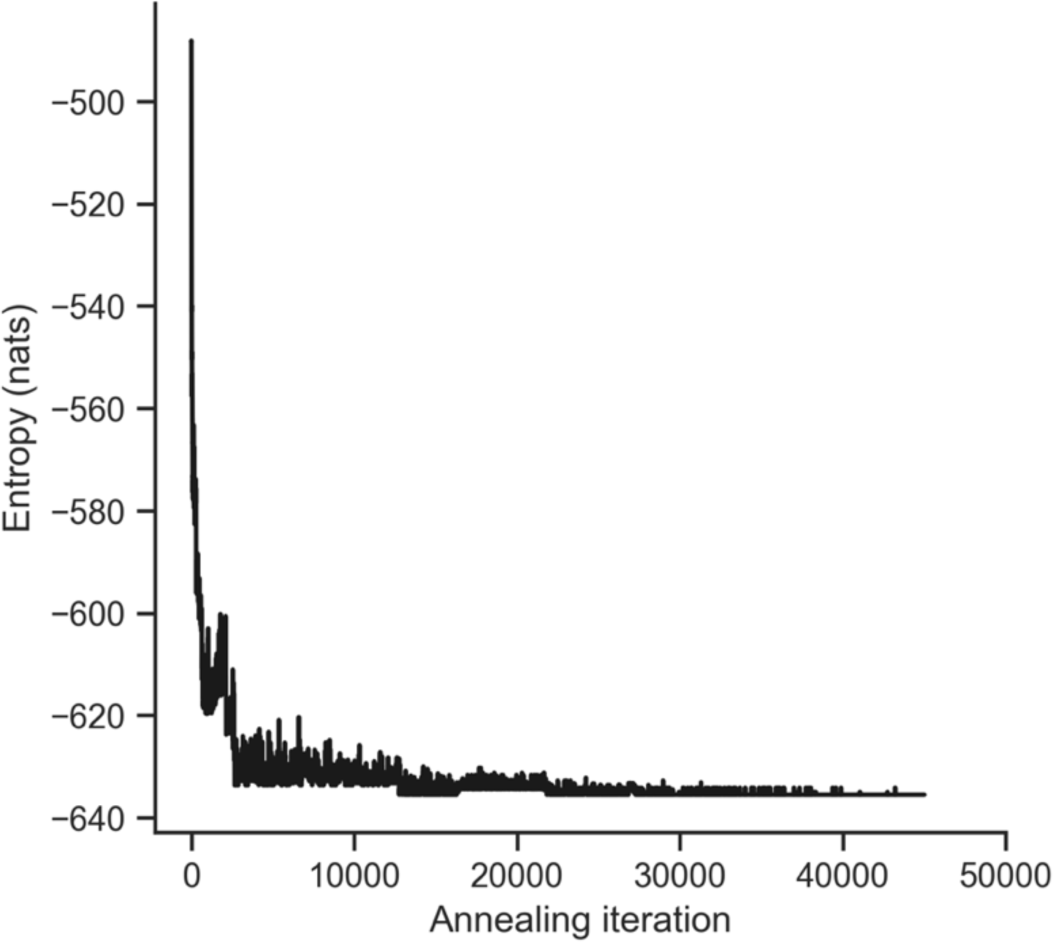
Line-plot of stochastic block model entropy with simulated annealing.

## References

1 Lived experience: shifting focus. Nature Mental Health 1, 145-145 (2023). 10.1038/s44220-023-00040-0

2 Mind. What is lived experience?, <https://www.mind.org.uk/workplace/influence-and-participation-toolkit/what/glossary-of-terms/#:~:text=What%20is%20lived%20experience%3F,experience%20of%20mental%20health%20problems.> (2024).

3 Drossman, D. A. Functional Gastrointestinal Disorders: History, Pathophysiology, Clinical Features and Rome IV. Gastroenterology (2016). 10.1053/j.gastro.2016.02.032

4 Drossman, D. A. & Hasler, W. L. Rome IV-Functional GI Disorders: Disorders of Gut-Brain Interaction. Gastroenterology 150, 1257–1261 (2016). 10.1053/j.gastro.2016.03.035

5 Burbige, E. J. Irritable bowel syndrome: diagnostic approaches in clinical practice. Clin Exp Gastroenterol 3, 127–137 (2010). 10.2147/CEG.S12596

6 Ruffle, J. K. et al. Constipation Predominant Irritable Bowel Syndrome and Functional Constipation Are Not Discrete Disorders: A Machine Learning Approach. The American journal of gastroenterology (2020). 10.14309/ajg.0000000000000816

7 Rajpurkar, P., Chen, E., Banerjee, O. & Topol, E. J. AI in health and medicine. Nature Medicine (2022). 10.1038/s41591-021-01614-0

8 Yiannakou, Y. et al. The PAC-SYM questionnaire for chronic constipation: defining the minimal important difference. Alimentary pharmacology & therapeutics 46, 1103–1111 (2017). 10.1111/apt.14349

9 Yiannakou, Y. et al. A randomized, double-blind, placebo-controlled, phase 3 trial to evaluate the efficacy, safety, and tolerability of prucalopride in men with chronic constipation. The American journal of gastroenterology 110, 741–748 (2015). 10.1038/ajg.2015.115

10 Emmanuel, A. et al. Factors affecting satisfaction with treatment in European women with chronic constipation: An internet survey. United European Gastroenterol J 1, 375–384 (2013). 10.1177/2050640613494200

11 Eltringham, M. T. et al. Functional defecation disorder as a clinical subgroup of chronic constipation: analysis of symptoms and physiological parameters. Scandinavian journal of gastroenterology 43, 262–269 (2008). 10.1080/00365520701686210

12 Palsson, O. S. & Whitehead, W. E. Psychological treatments in functional gastrointestinal disorders: a primer for the gastroenterologist. Clin Gastroenterol Hepatol 11, 208–216; quiz e222-203 (2013). 10.1016/j.cgh.2012.10.031

13 Ruffle, J. K., Farmer, A. D. & Aziz, Q. Artificial Intelligence Assisted Gastroenterology - Promises and Pitfalls. Am. J. Gastroenterol. 114, 422–428 (2019). 10.1038/s41395-018-0268-4.

14 Topol, E. The Topol Review: Preparing the healthcare workforce to deliver the digital future. (2019).

15 Topol, E. J. High-performance medicine: the convergence of human and artificial intelligence. Nat Med 25, 44–56 (2019). 10.1038/s41591-018-0300-7

16 Chang, L. & Heitkemper, M. M. Gender differences in irritable bowel syndrome. Gastroenterology 123, 1686–1701 (2002). 10.1053/gast.2002.36603

17 Newman, M. Networks. 2nd edn, (Oxford University Press, 2018).

18 Bullmore, E. & Sporns, O. Complex brain networks: graph theoretical analysis of structural and functional systems. Nat Rev Neurosci 10, 186–198 (2009). 10.1038/nrn2575

19 Barabasi, A. Network Science. 456 (Cambridge University Press, 2016).

20 Kleinberg, J. M. Authoritative Sources in a Hyperlinked Environment. J. Acm 46, 604– 632, numpages = 629 (1999). 10.1145/324133.324140

21 Goodfellow, I., Bengio, Y. & Courville, A. Deep Learning. (MIT Press, 2017).

22 Guthrie, E. et al. Cluster analysis of symptoms and health seeking behaviour differentiates subgroups of patients with severe irritable bowel syndrome. Gut 52, 1616–1622 (2003). 10.1136/gut.52.11.1616

23 Kettell, J., Jones, R. & Lydeard, S. Reasons for consultation in irritable bowel syndrome: symptoms and patient characteristics. The British journal of general practice : the journal of the Royal College of General Practitioners 42, 459–461 (1992).

24 Nelson, A., Herron, D., Rees, G. & Nachev, P. Predicting scheduled hospital attendance with artificial intelligence. npj Digital Medicine 2, 26 (2019). 10.1038/s41746-019-0103-3

25 Rubinov, M. & Sporns, O. Complex network measures of brain connectivity: uses and interpretations. Neuroimage 52, 1059–1069 (2010). 10.1016/j.neuroimage.2009.10.003

26 Siah, K. T., Wong, R. K. & Whitehead, W. E. Chronic Constipation and Constipation-Predominant IBS: Separate and Distinct Disorders or a Spectrum of Disease? Gastroenterol Hepatol (N Y) 12, 171–178 (2016).

27 Hill, A. B. The Environment and Disease: Association or Causation? Proc R Soc Med 58, 295–300 (1965).

28 Pearl, J. An introduction to causal inference. Int J Biostat 6, Article 7 (2010). 10.2202/1557-4679.1203

29 Lewis, S. J. & Heaton, K. W. Stool form scale as a useful guide to intestinal transit time. Scandinavian journal of gastroenterology 32, 920–924 (1997). 10.3109/00365529709011203

30 Marquis, P., De La Loge, C., Dubois, D., McDermott, A. & Chassany, O. Development and validation of the Patient Assessment of Constipation Quality of Life questionnaire. Scandinavian journal of gastroenterology 40, 540–551 (2005). 10.1080/00365520510012208

31 Herdman, M. et al. Development and preliminary testing of the new five-leve version of EQ-5D (EQ-5D-5 L). Quality of life research : an international journal of quality of life aspects of treatment, care and rehabilitation 20, 1727–1736 (2011). 10.1007/s11136-011-9903-x

32 Zhang, W. et al. Validity of the work productivity and activity impairment questionnaire--general health version in patients with rheumatoid arthritis. Arthritis Res Ther 12, R177 (2010). 10.1186/ar3141

33 Chen, T. & Guestrin, C. XGBoost: A Scalable Tree Boosting System. arXiv (2016). 10.48550/arXiv.1603.02754

34 Lundberg, S. & Lee, S.-I. A Unified Approach to Interpreting Model Predictions. arXiv (2017). 10.48550/arXiv.1705.07874

35 Peixoto, T. P. Nonparametric weighted stochastic block models. Physical Review E 97, 012306 (2018). 10.1103/PhysRevE.97.012306

36 Peixoto, T. P. Nonparametric Bayesian inference of the microcanonical stochastic block model. Physical Review E 95, 012317 (2017). 10.1103/PhysRevE.95.012317

37 Peixoto, T. P. Entropy of stochastic blockmodel ensembles. Physical Review E 85, 056122 (2012). 10.1103/PhysRevE.85.056122

38 Peixoto, T. P. in Advances in Network Clustering and Blockmodeling (eds Patrick. Doreian, Vladimir. Batagelj, & Anuška. Ferligoj) (John Wiley & Sons Ltd, 2019).

39 Ruffle, J. K. et al. Brain tumour genetic network signatures of survival. Brain 146, 4736–4754 (2023). 10.1093/brain/awad199

40 Cipolotti, L. et al. Graph lesion-deficit mapping of fluid intelligence. Brain 146, 167–181 (2022). 10.1101/2022.07.28.501722

41 Ruffle, J. K. et al. The autonomic brain: Multi-dimensional generative hierarchical modelling of the autonomic connectome. Cortex 143, 164–179 (2021). 10.1016/j.cortex.2021.06.012

## References

1 van Buuren, S. & Groothuis-Oudshoorn, K. mice: Multivariate Imputation by Chained Equations in R. 2011 45, 67 (2011). 10.18637/jss.v045.i03

2 Ruffle, J. K. et al. Brain tumour genetic network signatures of survival. Brain (2023).

3 Waskom, M. & Seaborn_Development_Team. seaborn. Zenodo (2020). 10.5281/zenodo.592845

4 Virtanen, P. et al. SciPy 1.0: fundamental algorithms for scientific computing in Python. Nature Methods 17, 261–272 (2020). 10.1038/s41592-019-0686-2

5 Barabasi, A. Network Science. 456 (Cambridge University Press, 2016).

6 Benjamini, Y. & Yekutieli, D. False Discovery Rate–Adjusted Multiple Confidence Intervals for Selected Parameters. Journal of the American Statistical Association 100, 71–81 (2005). 10.1198/016214504000001907

7 Ruffle, J. K., Farmer, A. D. & Aziz, Q. Artificial Intelligence Assisted Gastroenterology - Promises and Pitfalls. Am. J. Gastroenterol. 114, 422–428 (2019). 10.1038/s41395-018-0268-4.

8 Goodfellow, I., Bengio, Y. & Courville, A. Deep Learning. (MIT Press, 2017).

9 Chawla, N. V., Bowyer, K. W., Hall, L. O. & Kegelmeyer, W. P. SMOTE: Synthetic Minority Over-sampling Technique. *arXiv*, arXiv:1106.1813 (2011). 10.48550/arXiv.1106.1813

10 Chen, T. & Guestrin, C. XGBoost: A Scalable Tree Boosting System. arXiv (2016). 10.48550/arXiv.1603.02754

11 Lundberg, S. & Lee, S.-I. A Unified Approach to Interpreting Model Predictions. arXiv (2017). 10.48550/arXiv.1705.07874

12 Ruffle, J. K. et al. The autonomic brain: Multi-dimensional generative hierarchical modelling of the autonomic connectome. Cortex 143, 164–179 (2021). 10.1016/j.cortex.2021.06.012

13 Bullmore, E. & Sporns, O. Complex brain networks: graph theoretical analysis of structural and functional systems. Nat Rev Neurosci 10, 186–198 (2009). 10.1038/nrn2575

14 Peixoto, T. P. Nonparametric weighted stochastic block models. Physical Review E 97, 012306 (2018). 10.1103/PhysRevE.97.012306

15 Peixoto, T. P. Nonparametric Bayesian inference of the microcanonical stochastic block model. Physical Review E 95, 012317 (2017). 10.1103/PhysRevE.95.012317

57 Peixoto, T. P. in Advances in Network Clustering and Blockmodeling (eds Patrick. Doreian, Vladimir. Batagelj, & Anuška. Ferligoj) (John Wiley & Sons Ltd, 2019).

17 Peixoto, T. P. Descriptive vs. inferential community detection in networks: pitfalls, myths, and half-truths. arXiv (2021).

18 Newman, M. Networks. 2nd edn, (Oxford University Press, 2018).

19 Zalesky, A., Fornito, A. & Bullmore, E. T. Network-based statistic: identifying differences in brain networks. Neuroimage 53, 1197–1207 (2010). 10.1016/j.neuroimage.2010.06.041

20 Rubinov, M. & Sporns, O. Complex network measures of brain connectivity: uses and interpretations. Neuroimage 52, 1059–1069 (2010). 10.1016/j.neuroimage.2009.10.003

21 Peixoto, T. P. Hierarchical Block Structures and High-Resolution Model Selection in Large Networks. Physical Review X 4, 011047 (2014). 10.1103/PhysRevX.4.011047

22 Peixoto, T. P. Entropy of stochastic blockmodel ensembles. Physical Review E 85, 056122 (2012). 10.1103/PhysRevE.85.056122

23 Cipolotti, L. et al. Graph lesion-deficit mapping of fluid intelligence. Brain 146, 167–181 (2022). 10.1101/2022.07.28.501722

24 Peixoto, T. P. The graph-tool python library. figshare (2014). 10.6084/m9.figshare.1164194

25 Lemaitre, G., Nogueira, F. & Aridas, C., K. Imbalanced-learn: A Python Toolbox to Tackle the Curse of Imbalanced Datasets in Machine Learning. Journal of Machine Learning Research 18, 1–5 (2017).

26 Hunter, J. D. Matplotlib: A 2D Graphics Environment. Computing in Science & Engineering 9, 90–95 (2007). 10.1109/MCSE.2007.55

27 Harris, C. R. et al. Array programming with NumPy. Nature 585, 357–362 (2020). 10.1038/s41586-020-2649-2

28 Reback, J., McKinney, W. & jbrockmendel. pandas-dev/pandas: Pandas 1.0.3 (Version v1.0.3). *Zenodo* (2020). 10.5281/zenodo.3715232

29 Pedregosa, F., Varoquaux, G. & Gramfort, A. Scikit-learn: Machine Learning in Python. Journal of Machine Learning Research 12, 2825–2830 (2011).

